# Updated projections for COVID-19 omicron wave in Florida

**DOI:** 10.1101/2022.01.06.22268849

**Authors:** Thomas J. Hladish, Alexander N. Pillai, Ira M. Longini

## Abstract

In this report, we use a detailed simulation model to assess and project the COVID-19 epidemic in Florida. The model is a data-driven, stochastic, discrete-time, agent based model with an explicit representation of people and places. Using the model, we find that the omicron variant wave in Florida is likely to cause many more infections than occurred during the delta variant wave. Due to testing limitations and often mild symptoms, however, we anticipate that omicron infections will be underreported compared to delta. We project that reported cases of COVID-19 will continue to grow significantly and peak in early January 2022, and that the number of reported COVID-19 deaths due to omicron may be 1/3 of the total caused by the delta wave.

## Introduction

The first known case of the omicron variant of concern (VOC) of SARS-CoV-2 in Florida was reported on December 7, 2021 [1]. Since mid December, Florida has experienced extremely rapid spread of the omicron variant. For this report, we have revised our transmission model [2] to include a shorter time from exposure to becoming infectious (the *latent period*) for omicron compared to previous variants [3]. Coupled with being highly infectious relative to the delta variant, even among vaccinated people, assuming a shorter latent period allows our model to reproduce the rapid spread of omicron that has been observed. Important omicron-specific parameters are summarized in Table 1.

**Table 1:** Omicron parameter assumptions. Relative values are in comparison to the delta variant.

| Parameter | Assumed value |
| --- | --- |
| Relative transmission advantage | 2.0 |
| Relative severity | 0.25 |
| Relative latent period | 0.5 |
| Immune escape probability | 60% |

We calculate omicron’s transmission advantage as the ratio of the basic reproduction numbers of omicron and delta (i.e., 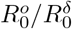). Omicron’s immune escape capability is modeled as a reduction in the probability that existing immune protection, whether infection- or vaccine-derived, will prevent infection. For comparison, we assume that delta’s immune escape probability is 15%. “Severity” is the probability that a person with symptoms will develop severe disease, a precondition we assume for hospitalization, ICU admission, or death. Similarly, we assume that vaccine efficacy against onward transmission (*V E*_*I*_) has also decreased (meaning vaccinated people are more able to transmit omicron compared to delta—see Table 2).

**Table 2:** Vaccine efficacy (*V E*) values assumed in our model, based on estimates from multiple Phase III trials and other published sources [8]. Delta *V E*_*S*_ values assume 15% immune escape. Omicron *V E*_*S*_ were calculated using Equation 1 with Wildtype as 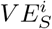 and are reported above for the lower and upper immune escape assumptions in Table 1. *V E*_*I*_ for omicron is also assumed to decrease. Other *V E* parameters for omicron will match those for other VOCs. *V E*_*S*_ and *V E*_*P*_ estimates have been revised since the October 08 report. Note on *V E* details: *V E*_*S*_ refers to vaccine efficacy against infection. *V E*_*P*_ refers to vaccine efficacy against symptoms given infection. *V E*_*H*_ refers to vaccine efficacy against severity given symptoms. *V E*_*I*_ refers to vaccine efficacy against onward transmission.

|  | Wildtype |  | Alpha |  | Delta |  | Omicron |  |
| --- | --- | --- | --- | --- | --- | --- | --- | --- |
|  | Dose 1 | Dose 2 | Dose 1 | Dose 2 | Dose 1 | Dose 2 | Dose 1 | Dose 2 |
| $VE_S$ | 0.4 | 0.8 | 0.4 | 0.8 | 0.34 | 0.68 | 0.4 | 0.8 |
| $VE_P$ | 0.67 | 0.75 | 0.67 | 0.75 | 0.67 | 0.75 | 0.67 | 0.75 |
| $VE_H$ | 0.9 | 1.0 | 0.9 | 1.0 | 0.9 | 1.0 | 0.45 | 0.9 |
| $VE_I$ | 0.4 | 0.8 | 0.4 | 0.8 | 0.4 | 0.8 | 0.2 | 0.4 |

Because of how contagious omicron is, and how rapidly the omicron wave is progressing, we do not believe dynamic changes to people’s personal-protective behaviors will play a major role in limiting transmission. Furthermore, as the state government has not indicated a plan to substantially change policies in response to omicron, we do not consider the possibility here.

Booster vaccine doses may substantially increase vaccine protection against disease caused by omicron [4]. At this time, 31.3% of Florida’s population has received a booster dose [5]. In these projections, we do not explicitly include the effects of boosting on omicron dynamics. Nonetheless, we recommend eligible people receive boosters, and we expect that an increase in booster uptake will result in more optimistic trajectories for the omicron wave in Florida.

## Results

We characterize the omicron epidemic wave using several metrics (Figure 1): simulated reported cases (yellow) are compared to empirical reported cases (black); simulated reported deaths (red) are compared to empirical reported deaths (black); simulated viral strain prevalence over time; total simulated infections (including asymptomatic infections and both reported and unreported cases); and time-varying reproduction number (*R*_*t*_) measured from the simulation. For the forecast period (December 2021 onward), we show 100 realizations of the model, with the mean trajectory overlaid as a bold line.

**Figure 1:**
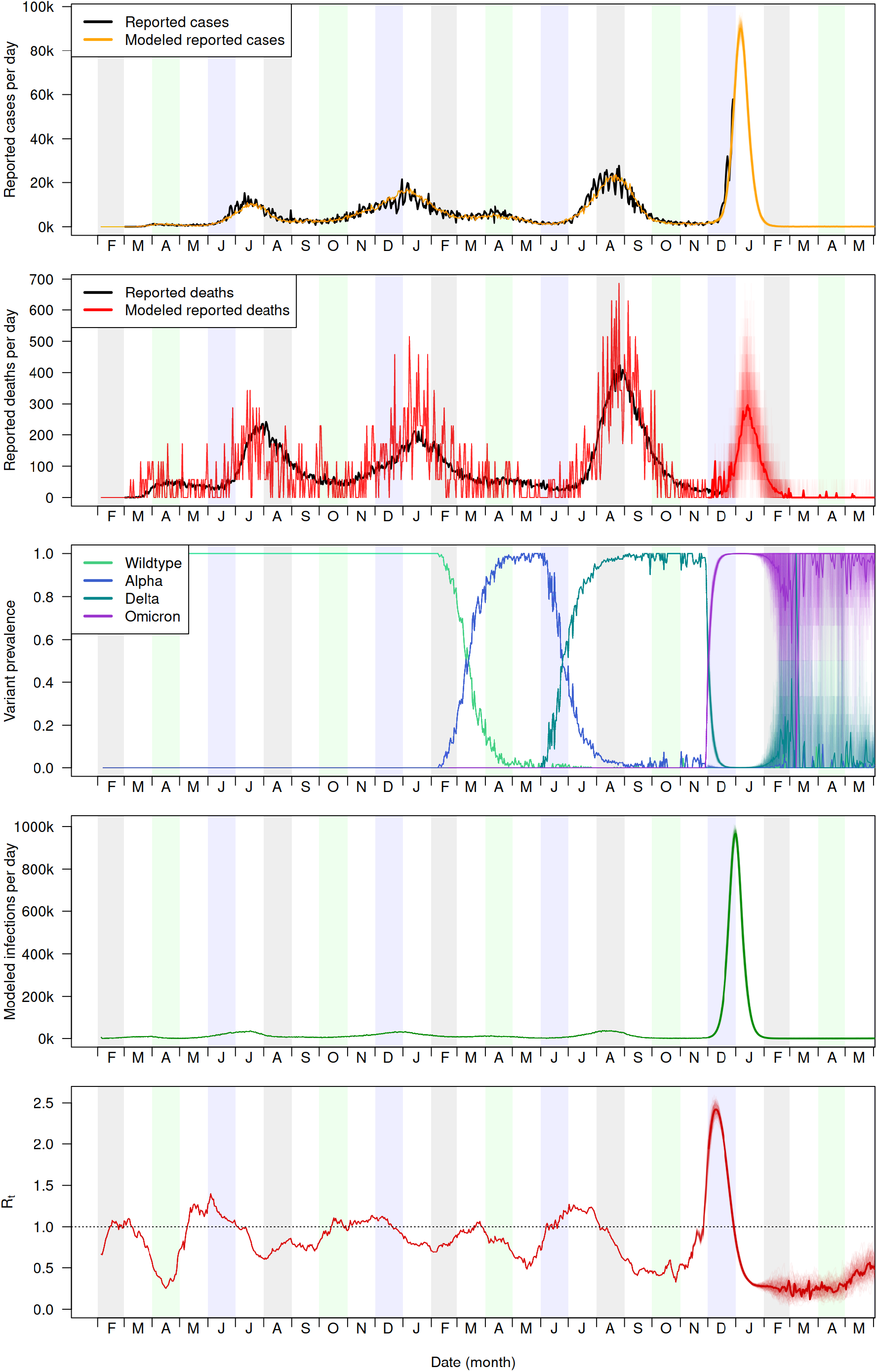
Projected reported cases, reported deaths, VOC prevalence, total infections, and reproduction number.

## Discussion

The omicron VOC is causing approximately twice the number of daily reported cases in Florida as were observed at the peak of the delta wave in late summer 2021. Using a detailed agent-based simulation model, we find that the omicron wave may cause an order of magnitude more infections than occurred during the delta wave, potentially infecting most of the state’s population in this wave alone. Preliminary data suggest that omicron infections may be less severe than those caused by delta, particularly among vaccinated people [6]. This means that despite causing more infections, it is possible that substantially fewer deaths will result from the omicron wave. We estimate that omicron will cause approximately 1/3 as many deaths as were caused by delta. Due to limitations in testing capacity, milder infections in vaccinees, and reduced sensitivity of some tests to the omicron variant, we believe that a smaller fraction of omicron infections will be detected as cases compared to previous waves. We expect reported cases to peak in the first half of January 2022.

Preliminary data suggest that boosting may dramatically increase protection against disease caused by omicron infections [4]. We therefore recommend eligible people receive boosters as soon as possible. Furthermore, absenteeism among infected health care workers combined with a rapidly increasing number of COVID-19 patients may substantially strain the health care system. This may be particularly serious for pediatrics, as children under 5 are not eligible for vaccination and may be more likely to experience severe omicron infections than adults [6]. We recommend particular care be taken to protect young children from exposure to the omicron variant.

## Methods

We have developed a detailed simulation model to serve as a tool for assessing the COVID-19 epidemic in Florida. The model is a data-driven, stochastic, discrete-time, agent based model with an explicit representation of people and places [7]. Households in the model are sampled from census and survey data in order to establish a realistic distribution of age, sex, comorbidity, employment and school-attendance status. Activities and interaction patterns affect how likely someone is to be exposed in the model, and age, health status, and healthcare seeking behavior affect how severe a person’s infection is likely to be. People go to work or school, visit friends, and patronize businesses in the model. The simulation includes closure of non-essential businesses, reduced school attendance, and changes in behaviors during the course of the pandemic. Our full Florida model represents 20.6 million people residing in 11.2 million households and 3.8 thousand long-term care facilities and who work in 2.3 million workplaces and attend 7.6 thousand schools. However, for this simulation study, we created a smaller, representative sample of the entire synthetic population totalling 375,000 people. We rescale the output from the model in order to estimate the cases and deaths for the entire state.

During each simulated day, infectious and susceptible individuals can aggregate in households, workplaces (both as employees and as customers), schools, long-term care facilities, and hospitals at different times in the day (Fig. 2). When susceptible and infectious people come together at the same location, there are new opportunities for the transmission of the virus.

**Figure 2:**
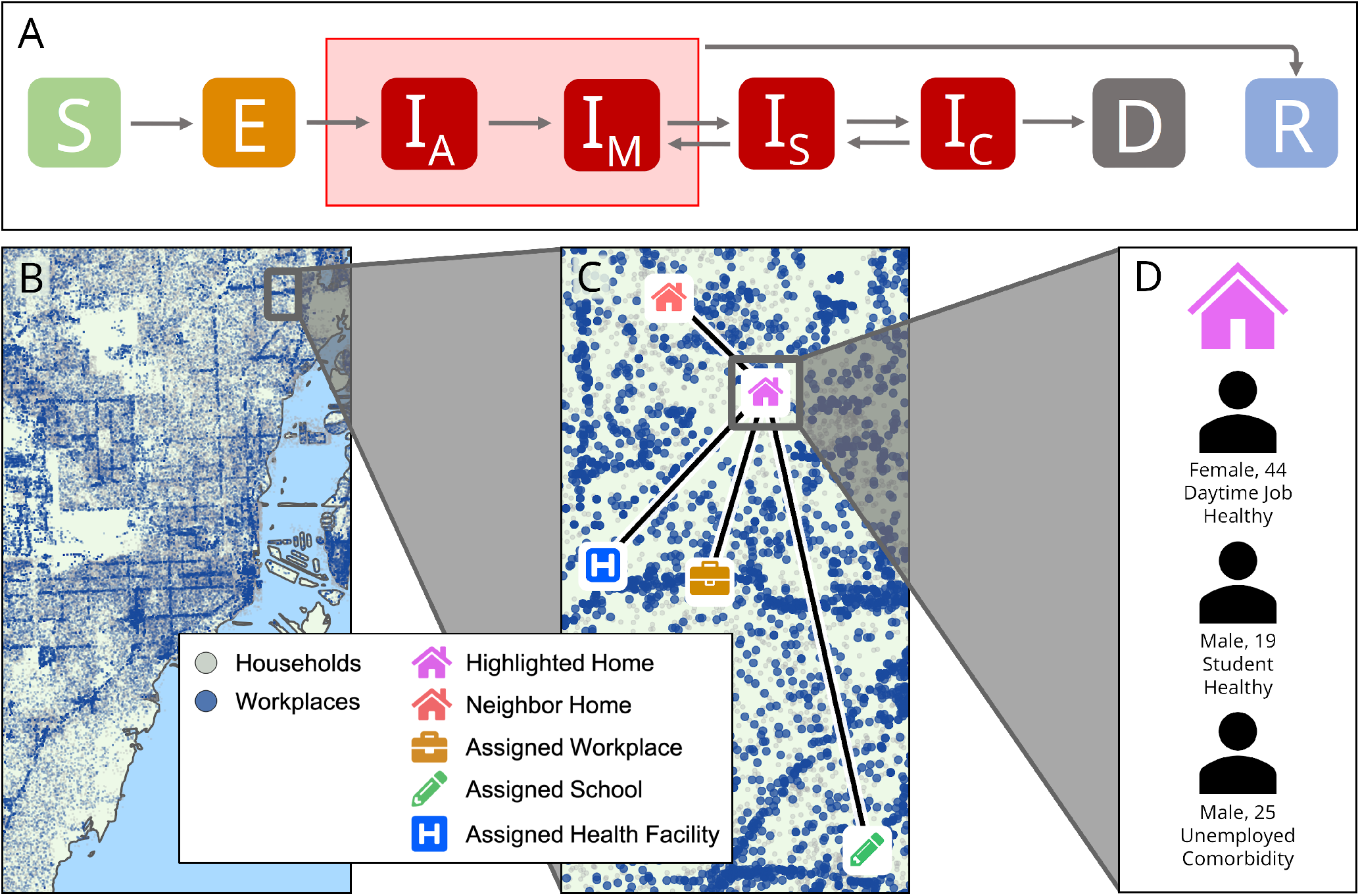
(A) Progression of the disease states in the model: susceptible (*S*) individuals may become exposed (*E*) to the virus, then progress to being infected (initially asymptomatic [*I*_*A*_], possibly progressing to mild [*I*_*M*_], severe [*I*_*S*_] or critical [*I*_*C*_]), and finally recovering (*R*) or dying (*D*). (B) Model locations of households and workplaces in an urban region (Miami, FL). (C) An example household. People may contact others by socializing with other households, by going to work or school, by going to the hospital, or by patronizing nearby businesses (not shown). (D) Attributes of the people in this household

If an individual becomes infected, the progression of the infection follows an *SEIRD* model where people progress through susceptible (*S*), exposed (*E*), infected (*I*), recovered (*R*), and dead (*D*) states. Additionally, infected individuals can develop mild (*I*_*A*_), severe (*I*_*M*_), or critical (*I*_*C*_) symptoms (Fig. 2. People who become ill can may seek healthcare, resulting in that individual receiving hospital care (for severe symptoms) or ICU care (for critical symptoms), which in turn lowers the risk of death.

Beyond non-pharmaceutical interventions (e.g. business or school closures, social distancing, stay-at-home orders), the model also represents vaccination of the synthetic population. In our model, we simulate a generalized mRNA vaccine (Table. 2) that performs similarly to the BioNTech and Moderna mRNA vaccines that have been used in Florida [8]. We simulate a rollout of vaccines that begins in January, 2021, with vaccine availability and campaign phases reflecting the vaccine rollout that has occurred in Florida (i.e. starting with healthcare workers and older members of the population and progressively widening eligibility to younger age groups).

We have revised our model of immunity to account for new data on immune dynamics and the effects of new variants. For vaccine-derived immunity, all people start with the same initial efficacy, whereas infections generate variable initial protection against reinfection. Both infection- and vaccine-derived immunity is modeled as leaky (in which every exposure has some chance of causing infection). We assume that efficacy against susceptibility (*V E*_*S*_) does not inherently wane, but does decrease due to changes in the circulating variants [9]. Efficacy against pathology (*V E*_*P*_) and against severe outcomes (*V E*_*H*_) remain constant over time. To calculate the current protection an individual has against infection due to vaccination (*V E*_*S*_) or infection (*IE*_*S*_), we use Equation 1 where 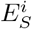 is the initial level of protection from either vaccination or infection, and Ω is the variant’s immune escape probability. In Table 2, we document our modeled vaccine efficacy values given the assumption that delta is a 15% immune escape mutant. Similar calculations are performed to determine a simulation’s *V E*_*S*_ or *IE*_*S*_ for omicron using immune escape assumptions.

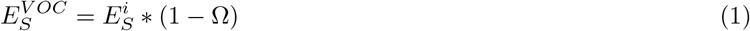

## Data Availability

All code used in the present study are available online at https://github.com/tjhladish/covid-abm
All data produced in the present study are available upon reasonable request to the authors

## Acknowledgements

This work was partially funded by NIH grants R01AI139761 and R56AI148284.

